# Gamma spectral event power is elevated in Fragile X Syndrome and associated with single trial gamma power during auditory chirp

**DOI:** 10.1101/2023.05.31.23290596

**Authors:** Yanchen Liu, Rui Liu, Paul S. Horn, Grace Westerkamp, Elizabeth Blank, Craig Erickson, Ernest V. Pedapati

**Author notes:** Corresponding author: Yanchen Liu, Division of Child and Adolescent Psychiatry, Division of Neurology, Cincinnati Children’s Hospital Medical Center, 3333 Burnet Ave, MLC 4002, Cincinnati, OH 45229.

## Abstract

**Background:** Fragile X syndrome (FXS) is a neurodevelopmental disorder resulting from silencing of the FMR1 gene. One of the most common and debilitating symptoms of FXS is sensory hyperarousal, especially in the auditory domain. Although the neural mechanisms of auditory hyperarousal in FXS are not well understood, electroencephalography (EEG) studies demonstrate increases in background gamma power during auditory paradigms, which are associated with more severe behavior and impairments in auditory synchronization.

**Methods:** High-frequency neural responses to the auditory chirp stimulus were studied in 36 individuals with FXS and 39 controls. Gamma Non-continuous high power events (spectral events) were quantified and compared from source localized EEG recordings. Correlation testing of spectral event properties was performed to averaged EEG features and clinical measures.

**Results:** Our results show that gamma event peak power was increased in the temporal source of male subjects with FXS (p<0.001, adj. p=0.008) as well as correlated with background average gamma power, while event number, event duration, and frequency span did not differ between groups. Further, absolute event power was positively correlated with clinical measures of obsessive behavior (R=0.63, adj. p=0.011) and stereotypic behavior (R=0.57, adj. p=0.031).

**Conclusions:** Our results indicate that gamma event peak absolute power likely underlies the increased background single trial gamma power observed during auditory processing in FXS, and that the temporal dynamics of gamma activity do not differ.

## Introduction

Fragile X syndrome (FXS) is an X-linked monogenetic neurodevelopmental disorder resulting from the loss of function of Fragile X messenger ribonuclear protein 1 (FMRP) (Richter and Zhao, 2021). Clinical presentation of FXS typically includes intellectual disability, autism spectrum disorders, high levels of anxiety, and heightened sensitivity to sensory stimuli (Salcedo-Arellano et al., 2020), with auditory hypersensitivity as one of the more common and debilitating symptoms in FXS (Rotschafer and Razak, 2014; Salvi et al., 2022). An amplitude-modulated broadband auditory chirp stimulus can be used to non-invasively probe cortical activity through electroencephalography (EEG) (Artieda et al., 2004).

Group-level differences from controls to the auditory chirp stimuli are robust and reproducible in individuals with FXS (Ethridge et al., 2019, 2017; Kenny et al., 2022). Specifically, FXS is associated with a reduction in phase-locking in the gamma band while asynchronous (background) gamma power is elevated compared to controls. This phenomenon appears to be conserved in Fmr1^-/-^ knockout mice (Fmr1^-/-^ KO) (Jonak et al., 2020). In humans, these gamma band elevations during the chirp have been correlated with behavior, neurocognitive function, and auditory attention (Ethridge et al., 2019, 2017). In addition, clinically-associated elevation of gamma power in FXS has been observed at rest (Pedapati et al., 2022; Wang et al., 2017) and during language tasks (Schmitt et al., 2020). Typically, gamma intertrial phase coherence (ITPC) and single trial power (STP) is calculated as the mean of 50 to 150 artifact-free event trials per participant. However, studying averaged gamma band power has significant limitations as they likely reflect a variety of brain processes (Buzsaki and Wang, 2012; Mably and Colgin, 2018) and lack specificity between major neuropsychiatric conditions (Newson and Thiagarajan, 2018).

Recent analysis of discontinuous activity at the trial level have provided new insights into the constituent components of mean power changes, underlying cognitive processes, and causal relationships leading to behavioral responses (Becker et al., 2020; Jones, 2016; Law et al., 2021; Vinding et al., 2020). Specifically, spectral events refer to discrete bursts of high-power activity within frequency bands which are otherwise obscured in averaged data (see **Figure 1A-B**). These events can be characterized based on their duration, peak power, frequency span, and quantity (see **Figure 1**). As the resulting event waveforms are related to the synchronized firing of neural populations, inferences about the underlying biophysical properties of the activity can be modeled and translated from animal models (Goswami et al., 2019; Neymotin et al., 2020). Thus, by analyzing the features of events at the individual trial level we can develop a finer grain characterization of neurodynamics within or between conditions.

**Figure 1 -.**
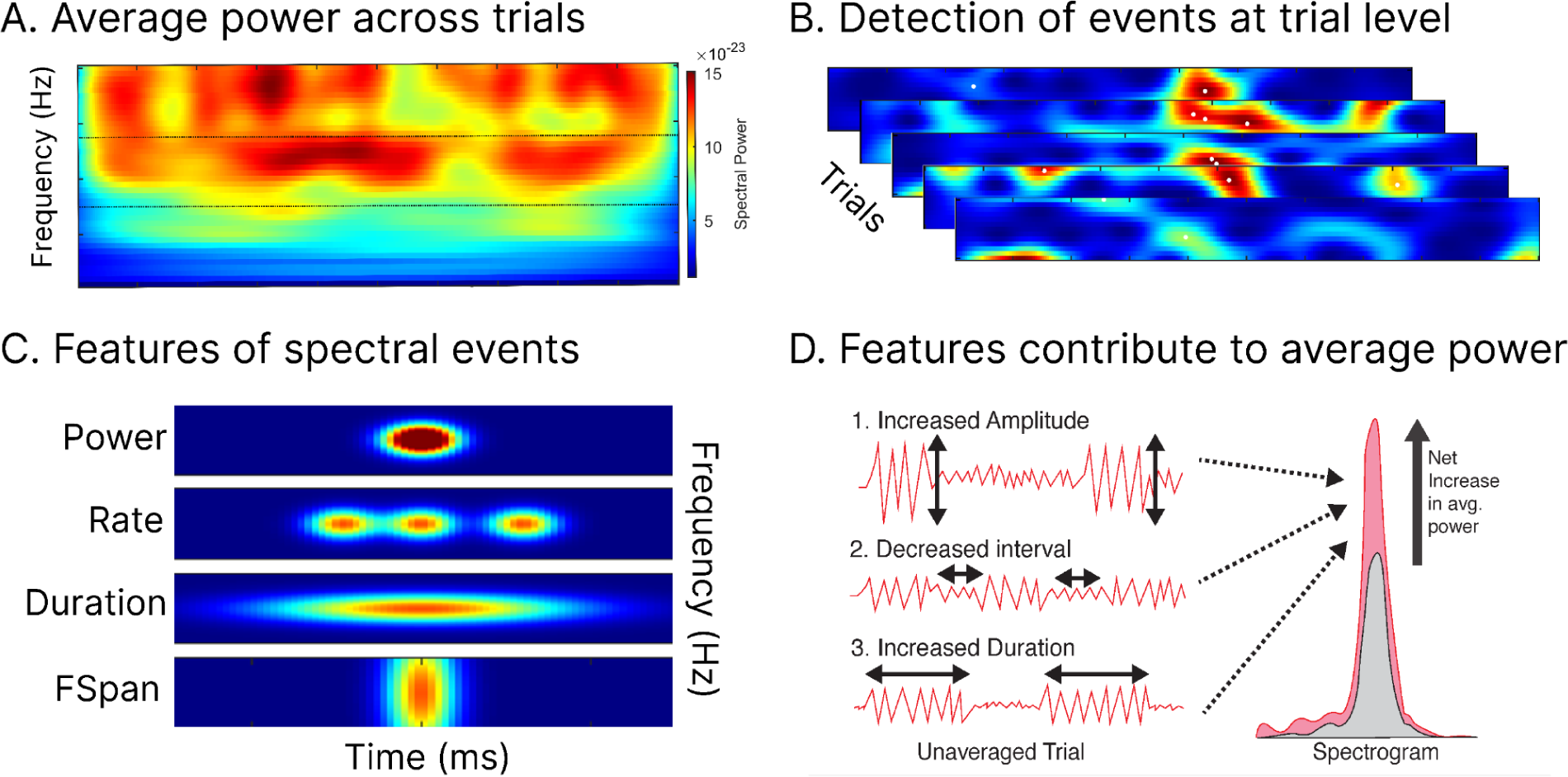
Spectral event features affect background power. **Figure 1. Gamma activity in individual trials appears in discrete high power bursts.** When the power spectra of many trials are averaged together, the trial level characteristics of these activity bursts are hidden. These features include event rate, peak power, duration, and frequency span. Changes to the features of spectral events can affect the average trial power. An increase in the rate, peak power, duration, or frequency span of spectral events would lead to an increase in the average power.

Herein, we parsed abnormalities in mean gamma power by studying gamma spectral events during the auditory chirp in a previously characterized, well-powered case-control cohort of individuals with FXS (Ethridge et al., 2019). We predicted that subjects with FXS would have a greater event rate, and increased event peak power compared to typically developing control (TDC) subjects, since existing research utilizing spectral events suggests that event rate and event power are most highly correlated with mean trial power (Shin et al., 2017). Because we observe an impairment of gamma phase locking and elevated asynchronous gamma power in FXS, we hypothesized that elevated gamma spectral event features would be inversely correlated with gamma ITC, and potentially represent decreased signal-to-noise in subjects with FXS that contributes to behavioral and auditory symptoms (Ethridge et al., 2019; Pedapati et al., 2022, Preprint).

## Methods

### EEG Recording

We analyzed event-related EEG recordings from an existing dataset of 36 participants with FXS (23 male and 13 female, mean age 25.4±10.3) and 39 age and sex matched typically developing control participants (22 male and 17 female, mean age 28.0±12.2) during an auditory chirp task (**see Table 1**). Scalp electrode analysis examining average participant responses of this EEG dataset was previously published (Ethridge et al., 2019). All participants provided written informed consent, and the study was approved by the institutional review board of Cincinnati Children’s Hospital Medical Center.

**Table 1 -.** Demographics and clinical variables

| Characteristic | FXS, N = 36<sup>1</sup> | TDC, N = 39<sup>1</sup> | p-value<sup>2</sup> |
| --- | --- | --- | --- |
| Sex |  |  |  |
| F | 13 (36%) | 17 (44%) |  |
| M | 23 (64%) | 22 (56%) |  |
| Deviation IQ | 42.398±29.081 | 102.950±8.349 | <0.001 |
| ADAMS General Anxiety | 7.700±4.764 | 2.441±2.608 | <0.001 |
| SCQ Total | 14.032±7.859 | 2.242±2.411 | <0.001 |
| ABC FXS subscale 1: irritability/aggression | 11.233±11.398 | 0.581±2.363 | <0.001 |
| ABC FXS subscale 4: Hyperactivity/Noncompliance | 7.700±5.932 | 0.935±2.081 | <0.001 |
| ABC FXS subscale 5: Inappropriate speech | 4.433±3.520 | 0.161±0.583 | <0.001 |
| ABC FXS subscale 2: lethargy/social withdrawal | 5.467±4.392 | 0.581±1.478 | <0.001 |
| ABC FXS subscale 3: stereotypy | 4.000±4.480 | 0.065±0.359 | <0.001 |
| WJ-III | 65.214±17.935 | 91.900±11.657 | <0.001 |
| Age at Visit | 25.399±10.274 | 27.975±12.189 | 0.5 |
| <sup>1</sup>n (%); Mean±SD |  |  |  |
| <sup>2</sup>Wilcoxon rank sum test; Wilcoxon rank sum exact test |  |  |  |

### Auditory Chirp Stimulus

Participants listened to 200 trials of an auditory chirp stimulus while watching silent video for compliance. The auditory chirp stimulus consists of pink noise that is amplitude modulated from 0 to 100 Hz over 2 seconds. The auditory chirp stimulus was presented using Sony MDR-V150 headphones at 65 dBa and tested to ensure no electrical artifact.

### Data acquisition and preprocessing

EEG data were collected at 1000 Hz using saline-based Hydrocel 128-channel EEG nets on a high-impedance NetAmp 400 amplifier (Magstim EGI, Eugene, Oregon). Impedance values were kept below 50 mOhms before recording. Before applying the EEG cap, Cz was identified and marked on participant’s heads as a guideline for net placement. All EEG data were blinded for preprocessing and analysis. Prior to analysis, raw recordings were bandpass filter filtered from 0.5 to 120 Hz and notch filtered at 60 Hz. Next, filtered data were visually inspected to remove noisy trials and interpolate channels containing excessive artifact. ICA decomposition was performed using the infomax algorithm in EEGLAB in order to remove independent components corresponding to eye, muscle, and channel artifacts (Delorme et al., 2012). Data were average referenced and epoched between −500 ms and 2750 ms. Spectral events were identified during the chirp stimulus period from 0 ms to 2750 ms.

### Source Localization

Artifact free data were source localized using a weighted minimum norm estimate (MNE) model in Brainstorm (Tadel et al., 2011). A depth weighting factor (order, .5; maximal amount, 10) was used to normalize amplitude values across sources (Lin et al., 2006). Default scalp electrode positions were co-registered to the Montreal Neurologic Institute ICBM152 common brain template (Lancaster et al., 2007). The forward model, or lead field matrix, was computed using Open MEEG resulting in a boundary element method head model that accounts for the brain, CSF, skin, and skull conductivity properties (Gramfort et al., 2010). The vertices were parcellated into 68 cortical nodes according to the Desikan-Killiany (DK) atlas (Desikan et al., 2006). To minimize variability in source localization across individuals, spectral event features for 68 DK atlas nodes were averaged into 14 cortical and subcortical regions (**see Supplemental Figure 1**) (Left frontal LF, right frontal RF, left temporal LT, right temporal RT, left parietal LP, right parietal RP, left prefrontal LPF, right prefrontal RPF, left occipital LO, right occipital RO, left cingulate LL, right cingulate RL, left central LC, and right central RC).

**Table 2 -.** Event feature significant group contrasts

| **Sex** | **Region** | **Feature** | **n (FXS)** | **n (TDC)** | **Statistic** | **p** | **p.adj** |
| --- | --- | --- | --- | --- | --- | --- | --- |
| M | LT | Event power absolute | 23 | 22 | 418 | 0.0001 | 0.008 |
| M | RT | Event power absolute | 23 | 22 | 391 | 0.0014 | 0.054 |
| F | LF | Event fspan | 13 | 17 | 172 | 0.0091 | 0.202 |
| M | LO | Event power absolute | 23 | 22 | 362 | 0.0127 | 0.202 |
| F | LF | Event power normalized | 13 | 17 | 169 | 0.0136 | 0.202 |
| F | RO | Event duration | 13 | 17 | 167 | 0.0174 | 0.202 |
| M | RP | Event power absolute | 23 | 22 | 357 | 0.0177 | 0.202 |

### Identification of Gamma Spectral Events

Spectral events in the gamma frequency band were extracted from source-localized EEG time series using the Spectral Events MATLAB Toolbox (Shin et al., 2017) (https://github.com/jonescompneurolab/SpectralEvents). Gamma frequency was defined as 30 to 55 Hz. The algorithm for detecting spectral events identifies frequency specific activity that surpasses an empirically-derived threshold (**see Figure 2**). To determine the threshold for calculating spectral events, a range of power thresholds between 1 and 20 factors of median (FOM) were used to calculate spectral events. The area above the threshold was correlated with mean trial power at each threshold to determine the threshold value at which area above threshold best correlates with mean trial power. In this study, it was determined that a power threshold of approximately 4 factors of median would best capture spectral events in the gamma frequency band (See **Figure 2**). Once the threshold was established, spectral events were identified by thresholding the time-frequency representation (TFR) within the gamma band above the FOM cutoff value. Local maxima were then identified in each suprathreshold area, and peaks with the highest power in each suprathreshold area were selected. This process identifies discrete bursts of high-power activity within the gamma frequency band, whose features were analyzed to gain insights into the underlying neurodynamics of auditory processing in individuals with FXS.

**Figure 2 -.**
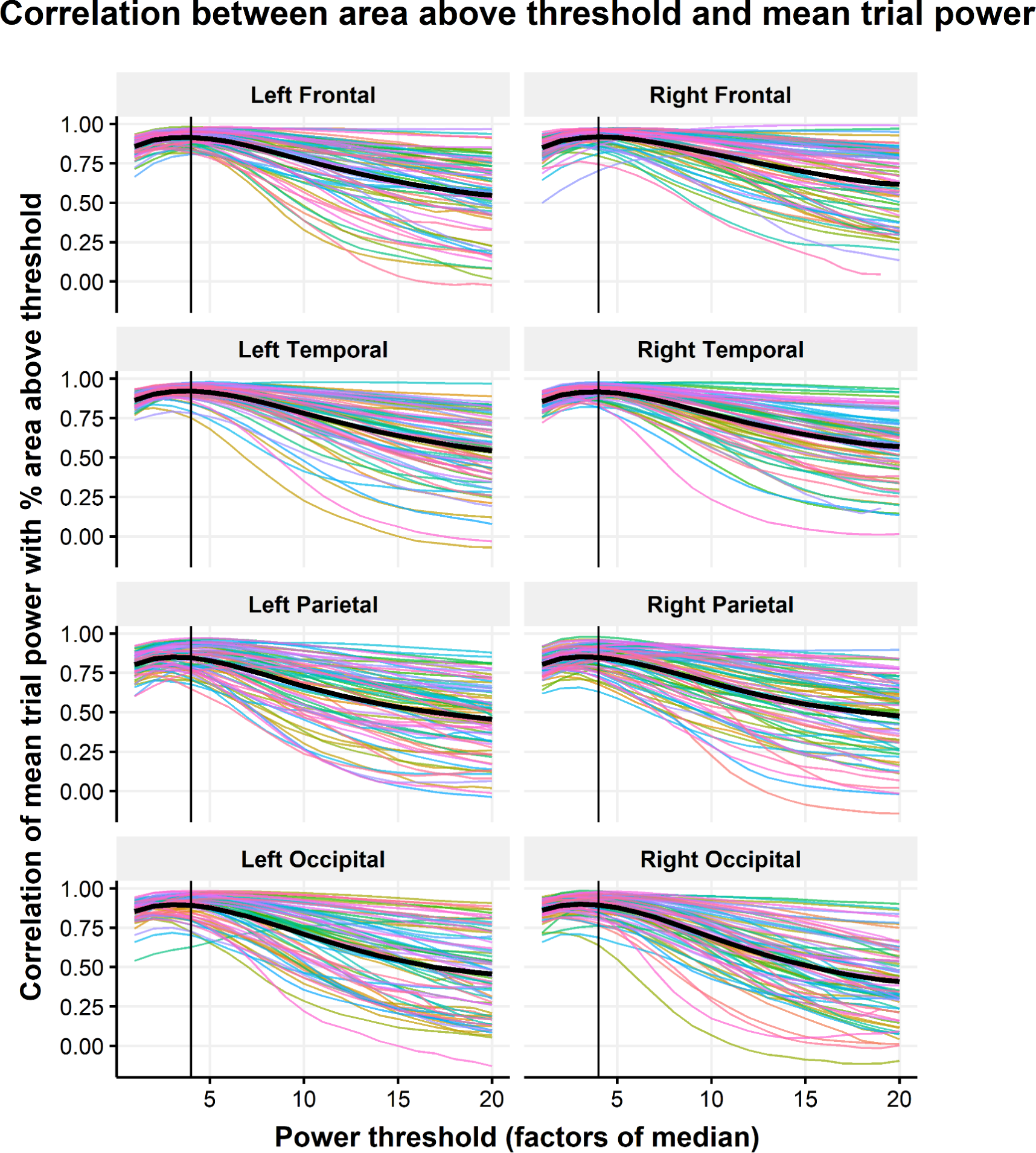
Determination of FOM threshold for gamma events. **Figure 2. A threshold of 4x median power was used to identify gamma spectral events.** The correlation across trials between mean trial power and area above threshold was calculated for a series of thresholds ranging from 1 to 20 Hz. Abbreviations: LF, Left Frontal; RF, Right Frontal; LO, Left Occipital; RO, Right Occipital; LT, Left Temporal; RT, Right Temporal

### Features associated with Gamma Spectral Events

We calculated five separate features of spectral events: event rate, event duration, absolute event power, normalized event power, and 5) event frequency span (Fspan). Event Fspan is defined as the highest frequency minus the lowest frequency for a given event. Normalized event power refers to the power at the peak of the spectral events normalized by the median power across trials at each frequency, in units of factors of median, while absolute event power was calculated by extracting the power value at the peak of each spectral event without normalizing to median power across trials. Event features were calculated across the entire chirp trial, and averaged over trials for each channel for the group differences analysis. When calculating averages for normalized event power, absolute event power, event duration, and event frequency, only trials that contained at least one event were used. For event rate, all trials were taken into account when calculating average rate across trials, including trials with zero events.

### Statistical Analysis

Group comparisons of spectral event features were calculated between FXS and TDC male subjects, and between FXS and TDC female subjects using wilcoxon-ranked sum tests. A 5% FDR correction was applied to adjust for multiple comparisons (see **Table 3, Figure 3**).

**Figure 3 -.**
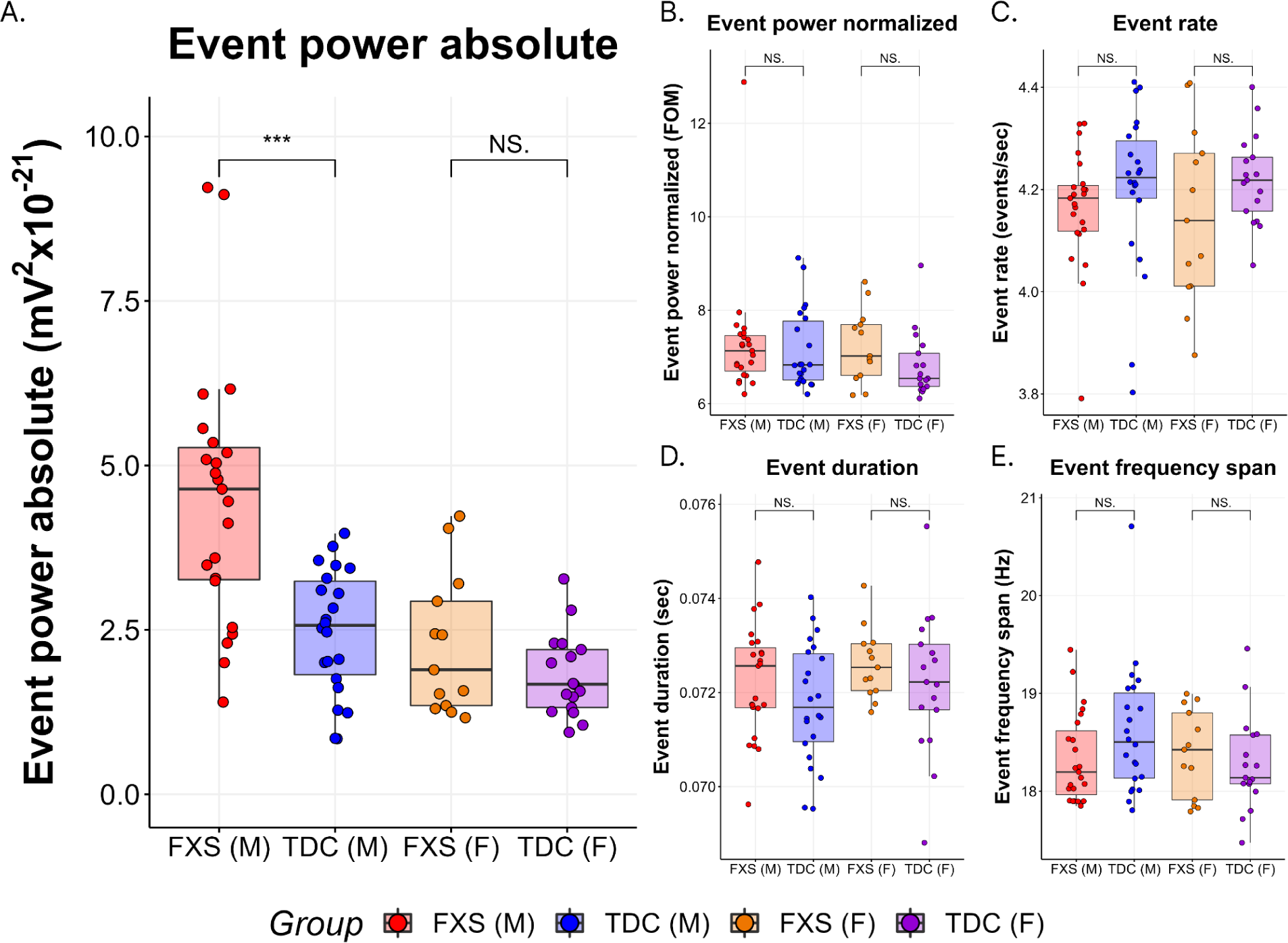
Mean event peak power increased in FXS left temporal. **Figure 3. Event power absolute is increased in the left temporal source of male FXS subjects.** Male subjects with FXS have increased absolute event power in the left temporal source compared to male TDC subjects (W=418, p<0.0001, adj. p=0.0075). No other event features were significantly different in FXS and TDC, when comparing across sex (see **Table**.

**Table 3 -.**
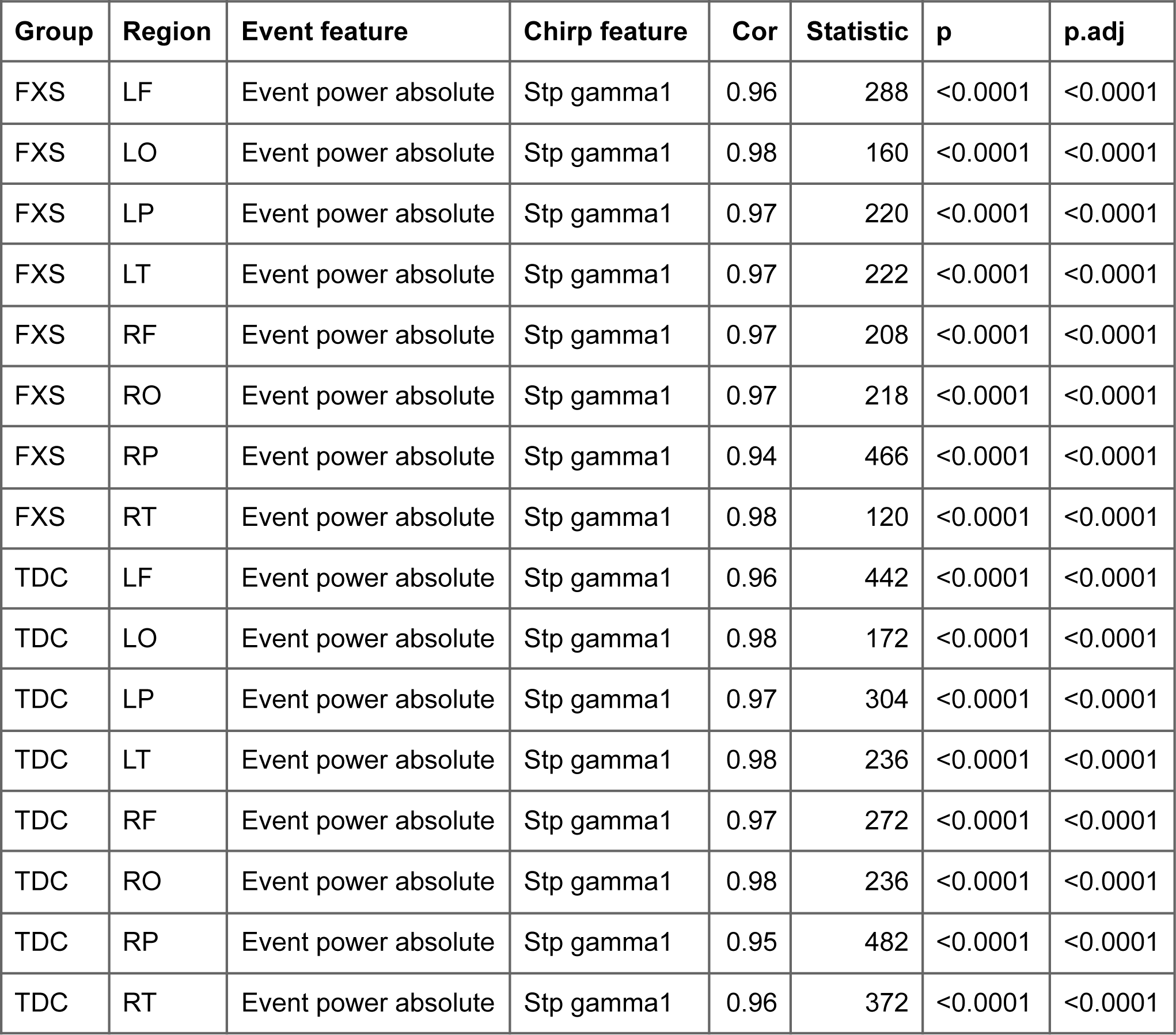
Subject level correlations between absolute event power and gamma STP

| Group | Region | Event feature | Chirp feature | Cor | Statistic | p | p.adj |
| --- | --- | --- | --- | --- | --- | --- | --- |
| FXS | LF | Event power absolute | Stp gamma1 | 0.96 | 288 | <0.0001 | <0.0001 |
| FXS | LO | Event power absolute | Stp gamma1 | 0.98 | 160 | <0.0001 | <0.0001 |
| FXS | LP | Event power absolute | Stp gamma1 | 0.97 | 220 | <0.0001 | <0.0001 |
| FXS | LT | Event power absolute | Stp gamma1 | 0.97 | 222 | <0.0001 | <0.0001 |
| FXS | RF | Event power absolute | Stp gamma1 | 0.97 | 208 | <0.0001 | <0.0001 |
| FXS | RO | Event power absolute | Stp gamma1 | 0.97 | 218 | <0.0001 | <0.0001 |
| FXS | RP | Event power absolute | Stp gamma1 | 0.94 | 466 | <0.0001 | <0.0001 |
| FXS | RT | Event power absolute | Stp gamma1 | 0.98 | 120 | <0.0001 | <0.0001 |
| TDC | LF | Event power absolute | Stp gamma1 | 0.96 | 442 | <0.0001 | <0.0001 |
| TDC | LO | Event power absolute | Stp gamma1 | 0.98 | 172 | <0.0001 | <0.0001 |
| TDC | LP | Event power absolute | Stp gamma1 | 0.97 | 304 | <0.0001 | <0.0001 |
| TDC | LT | Event power absolute | Stp gamma1 | 0.98 | 236 | <0.0001 | <0.0001 |
| TDC | RF | Event power absolute | Stp gamma1 | 0.97 | 272 | <0.0001 | <0.0001 |
| TDC | RO | Event power absolute | Stp gamma1 | 0.98 | 236 | <0.0001 | <0.0001 |
| TDC | RP | Event power absolute | Stp gamma1 | 0.95 | 482 | <0.0001 | <0.0001 |
| TDC | RT | Event power absolute | Stp gamma1 | 0.96 | 372 | <0.0001 | <0.0001 |

To determine the association between spectral event features and mean trial Gamma power, correlations were performed using Spearman’s rho (see **Supplemental Table 1, Figure 4**).Correlations across trials between event features and trial power were performed per DK atlas source node within each subject, and averaged across nodes into cortical regions. Kruskal-Wallis H tests were used to compare correlation values across the 4 diagnostic subgroups (groups and sex), for each cortical region and spectral event feature (see Supplemental Table 1, Figure 4).

**Figure 4 -.**
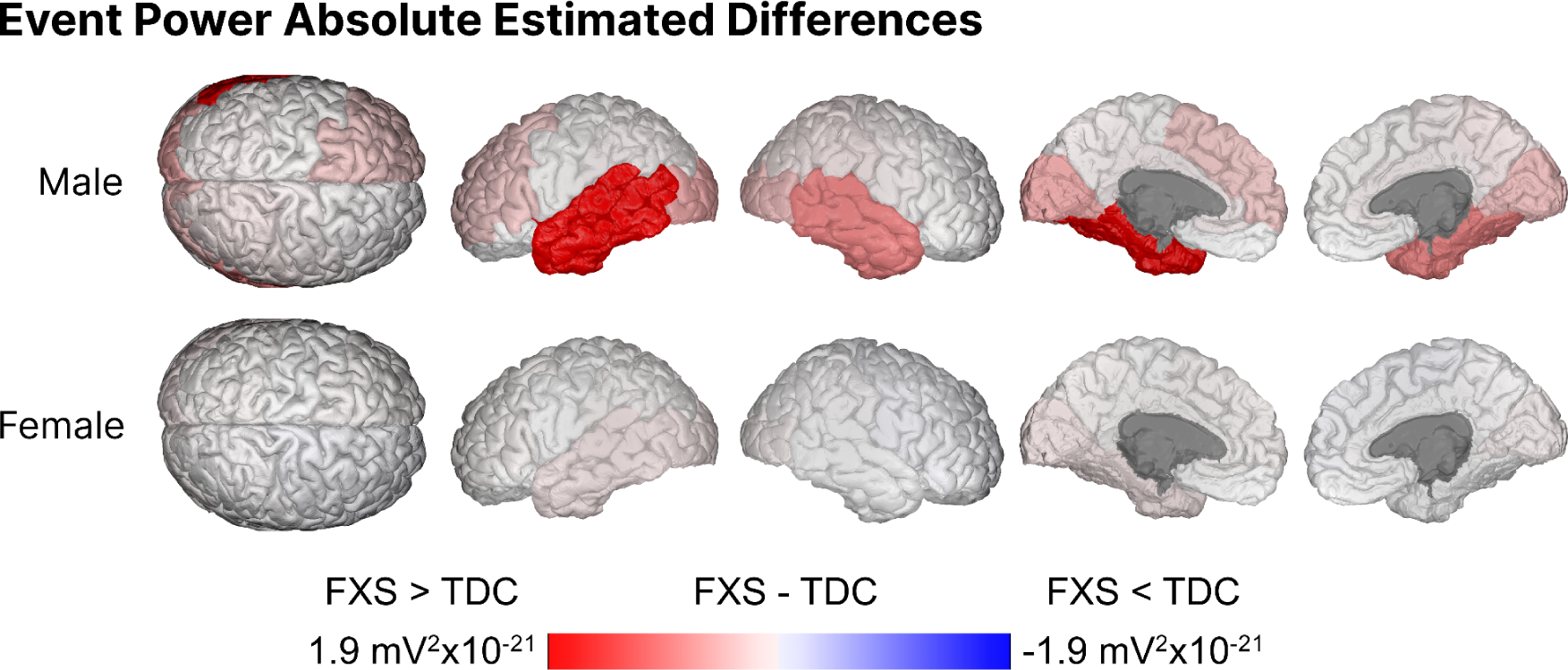
Event power absolute estimated differences across regions. **Figure 3. Event power absolute is increased in the left temporal source of male FXS subjects.** Subjects with FXS have marginally increased absolute event power in the right temporal source after correction (W=391, p=0.00136, adj. p=0.0544).

Average event rate over time was calculated using a sliding window over the duration of the trial with a 100 ms window in 10 ms steps across the duration of the chirp stimulus (see **Figure 5**). Point wise t tests were calculated to determine whether average event rate differed across diagnostic groups and sex over the duration of the trial, with a 5% FDR correction applied for each time point. To avoid edge effects, only time points from −250 to 2500 ms are selected for the event feature over time sliding window analysis.

**Figure 5 -.**
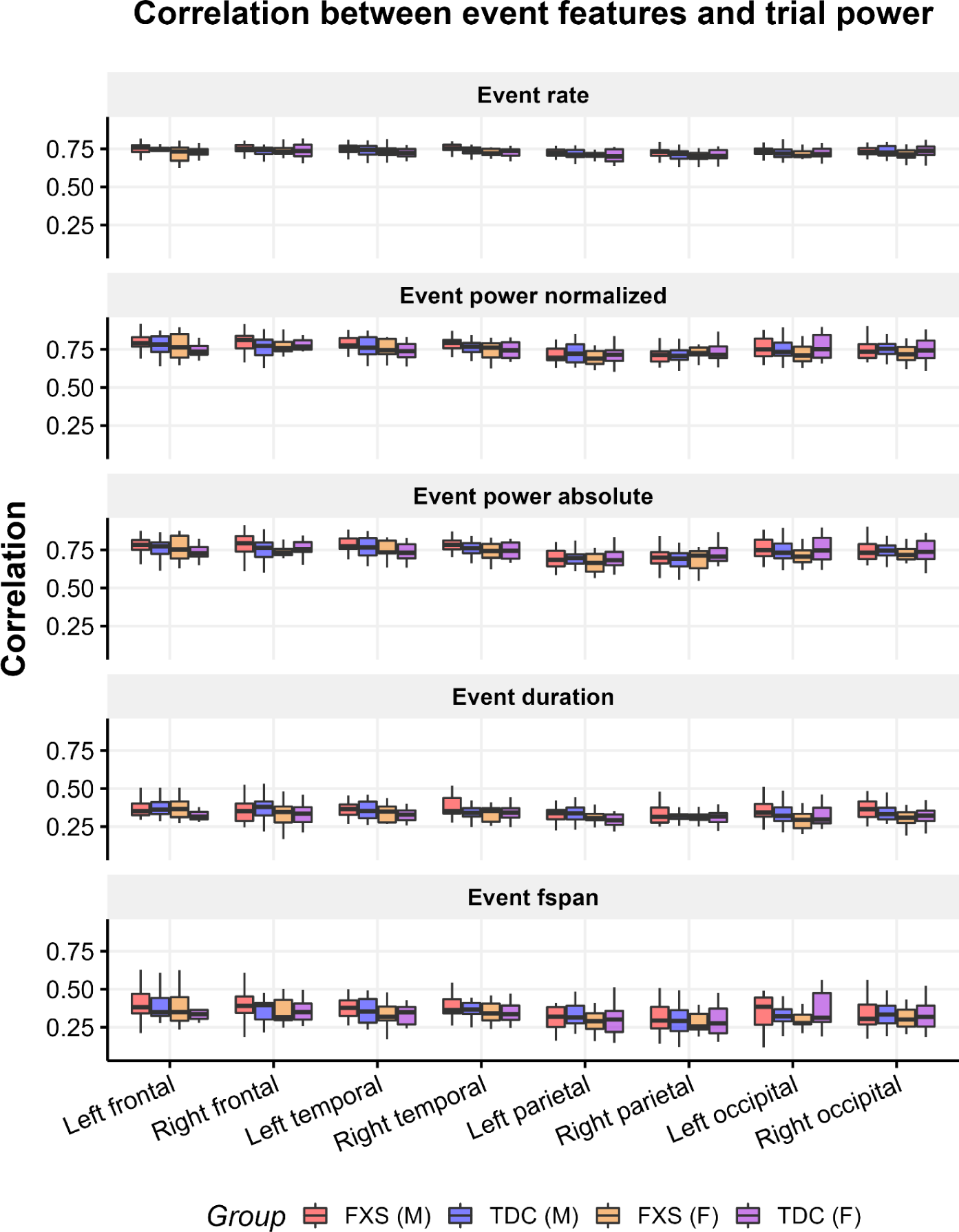
Event features contribute similarly to trial power across groups. **4. Event features contribute similarly to trial power across groups.** Trial average spectral event rate, average event power, average event duration, and average event frequency span were correlated with average trial power in order to determine which features contribute most to average gamma power. Correlation values did not differ significantly across group and sex for any region or event feature after correction (see **Supplemental Table 1**), suggesting that event features contribute similarly to event power across groups.

Correlations across subjects between spectral event features and clinical variables as well as gamma ITC were performed using Spearman’s rank correlations. The region of interest used for clinical correlations was chosen to be the left temporal region, as the differences in absolute event peak power were strongest in the left temporal region. A 5% FDR correction was applied to adjust for multiple comparisons. Correlations were performed between spectral event features and gamma ITC because existing research indicates that gamma STP is negatively associated with ITC (Ethridge et al., 2017, Ethridge et al., 2019)

## Results

We observed significant differences in gamma absolute peak power between FXS and TDC participants in the temporal source. Furthermore, gamma peak power was positively correlated with gamma STP across all subjects, suggesting that peak power contributes to average trial power. However, average gamma event rate, duration, and frequency in FXS was not different compared to TDC. Gamma event power was significantly correlated with ADAMS Anxiety and ABC obsessive compulsive behavior assessments.

### Gamma event peak power increased in FXS temporal lobe

Gamma event absolute power was significantly increased in male subjects with FXS compared to TDC in the left temporal source (W=418, p<0.0001, adj. p=0.0075). Increased absolute event power is also present in the right temporal source, though it does not survive correction (W=391, p=0.00136, adj. p=0.0544) (see **Figure 3, 4**). Gamma event absolute peak power did not differ significantly across groups in other regions (see **Table 3**).

Furthermore, gamma event absolute peak power and gamma single trial power were significantly correlated across all cortical sources (see **Table 3**). Other event features were not significantly correlated with gamma single trial power.

### Event features contribute similarly to event power across diagnostic group and sex

To assess whether event features contribute to trial power similarly across group and sex, event rate, normalized power, duration, and frequency span were correlated with normalized trial power (see **Figure 2**). There were no regions or features where correlation between event feature and mean trial power differed across diagnostic groups and sex (see **Supplemental Table 1**).

### Absolute event power associated with greater impairment in clinical measures

Gamma event power in the left temporal source was significantly correlated with ADAMS obsessive compulsive behavior scale score (r(31)=.633, p=.000132, adj. p=.00858) and with the ABC FXS subscale 3: stereotypy (r(30)=.568, p=.00106, adj. p=.0344) (see **Figure 5**, **Table 5**).

Gamma event absolute power was negatively correlated with gamma ITC for TDC subjects (r(39)=-.50, p=0.0014, adj. p=0.03) (see **Table 4**, **Figure 8**), which aligns with previous scalp level findings showing negative correlations between gamma STP and gamma ITC (Ethridge et al., 2017, Ethridge et al., 2019). However, for subjects with FXS, there was no significant correlation between event power absolute and gamma ITC (r(36)=-.08, p=0.644, adj. p=0.93).

**Figure 6 -.**
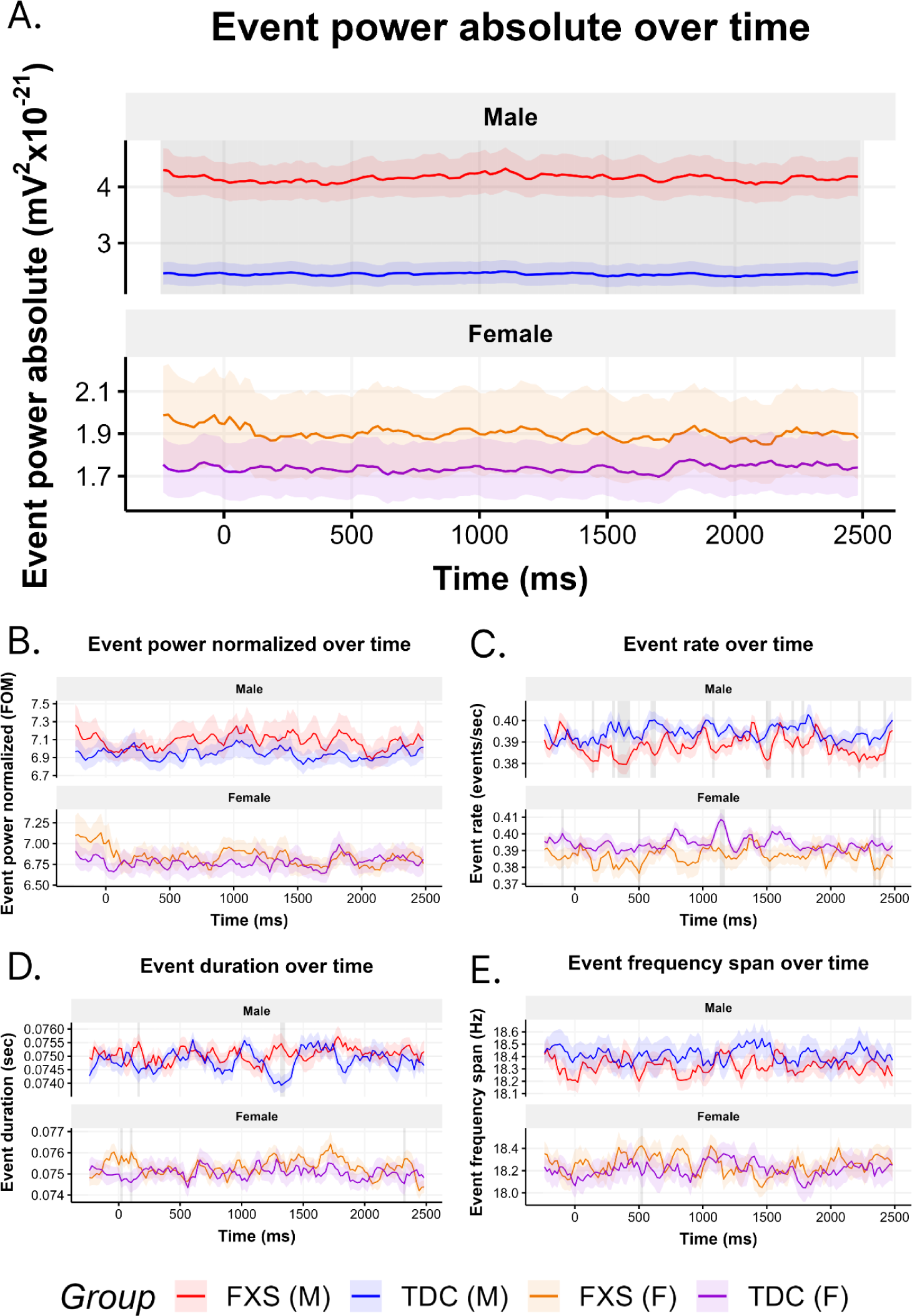
Event rate over time does not differ across groups. **Figure 4A. Gamma event rate does not change significantly over the course of trial or differ across groups.** A sliding window analysis was performed to calculate the average event rate for each subject using a 100 ms window in 10 ms steps across the duration of the chirp stimulus. Point wise Wilcoxan ranked sum tests were used to determine if event rate at given points in time differed across groups, with time points where event rate differed marked in grey.

**Table 4 -.** Subject level correlations between spectral event features and gamma ITC in left temporal source

| Group | Region | Event feature | Chirp feature | Cor | Statistic | p | p.adj |
| --- | --- | --- | --- | --- | --- | --- | --- |
| FXS | LT | Event rate | ltc40 | 0.01 | 7,714 | 0.967 | 1. |
| TDC | LT | Event rate | ltc40 | 0.12 | 8,710 | 0.471 | 0.86 |
| FXS | LT | Event power absolute | ltc40 | -0.08 | 8,388 | 0.644 | 0.93 |
| TDC | LT | Event power absolute | ltc40 | -0.50 | 14,800 | 0.0014 | 0.03 |
| FXS | LT | Event power normalized | ltc40 | -0.01 | 7,878 | 0.936 | 0.99 |
| TDC | LT | Event power normalized | ltc40 | -0.06 | 10,482 | 0.712 | 0.95 |
| FXS | LT | Event duration | ltc40 | 0.17 | 6,476 | 0.33 | 0.81 |
| TDC | LT | Event duration | ltc40 | 0.01 | 9,796 | 0.959 | 0.99 |
| FXS | LT | Event fspan | ltc40 | 0.15 | 6,606 | 0.382 | 0.82 |
| TDC | LT | Event fspan | ltc40 | -0.23 | 12,124 | 0.164 | 0.66 |

## Discussion

While elevated background gamma power during auditory chirp is a well-established characteristic of FXS, the underlying patterns of gamma activity that lead to this increase are unclear. Analyzing EEG power using discrete high power spectral events provides insight into the trial level patterns in gamma activity that are hidden in typical analyses of EEG band power.

Our main findings include 1) male subjects with FXS have increased gamma event absolute peak power temporal sources compared to TDC, 2) other event features do not differ across diagnostic groups, 3) absolute event power is strongly associated with gamma single trial power across subjects, and 4) increased absolute event power in subjects with FXS is associated with clinical measures of obsessive compulsive and stereotypic behavior. These results shed light on the temporal dynamics of asynchronous gamma power in fragile x syndrome.

### Event power absolute is related to subject level variation in average gamma power

It was hypothesized that differences in gamma event features would be localized to temporal regions, as existing source localized findings suggest that gamma STP is most increased in temporal regions (Pedapati et al., 2022, Preprint). The observed increase in gamma event peak power in the temporal source in FXS (see **Figure 2**), and strong correlations between absolute event peak power and gamma STP (see **Figure 7**) suggests that peak event power contributes to the subject level variation in gamma single trial power. Importantly, no other event features were significantly correlated with gamma single trial power across subjects.

**Figure 7 -.**
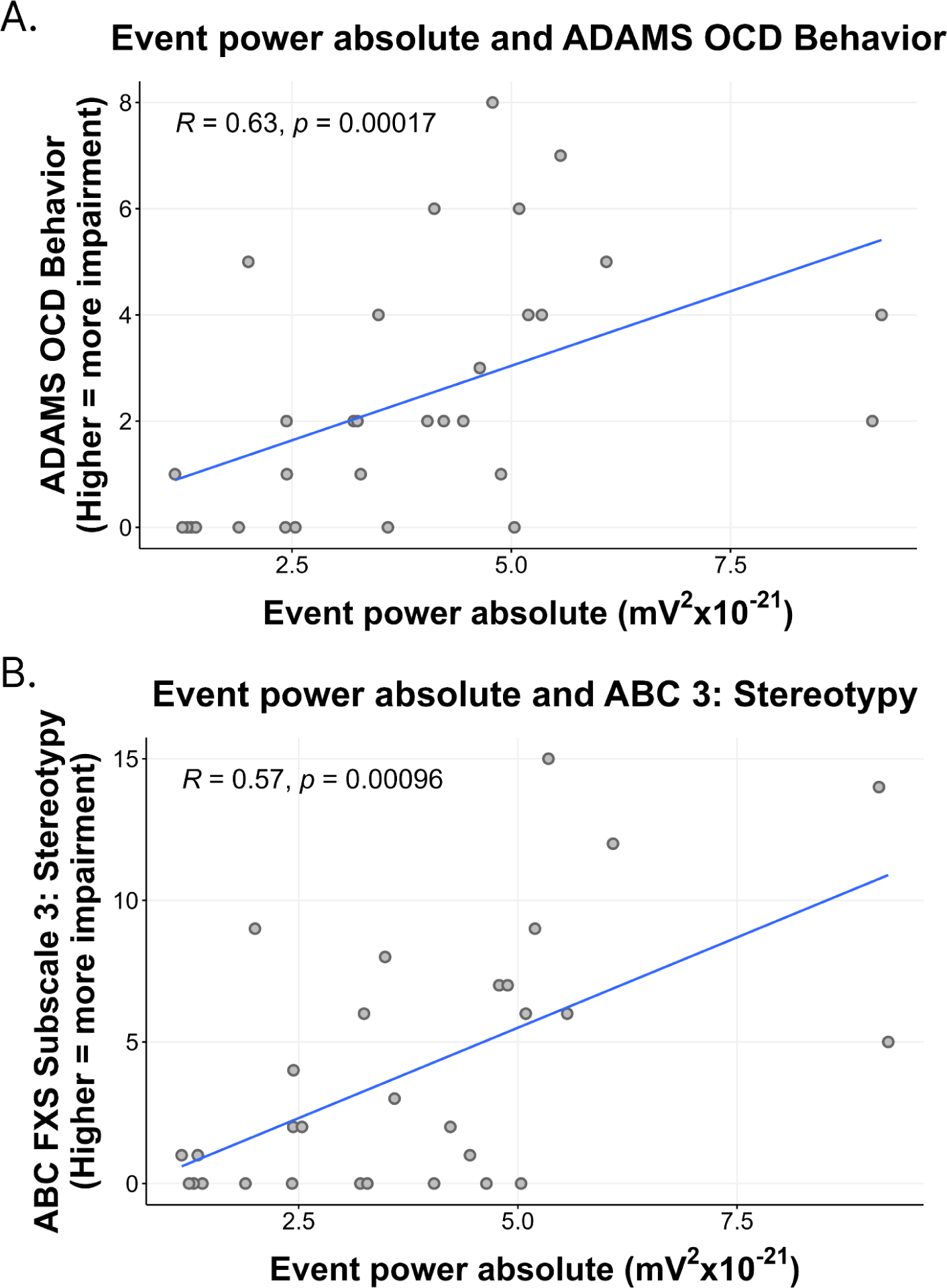
Event peak power is positively correlated with FXS severity. **Figure 4. Clinical measure correlations.** Gamma event power in the left temporal source was significantly correlated with ADAMS obsessive compulsive behavior scale score (r(31)=.63, p=.0002, adj. p=.011) and with the ABC FXS subscale 3: stereotypy (r(30)=.57, p=.0010, adj. p=.031).

### No difference in other features of gamma events between FXS and TDC

While significant It is unclear whether differences in peak power are directly responsible for an increase in average power. Importantly, no group level differences in event rate, duration, frequency span, or normalized power were found. One explanation for this finding is that the amount of gamma activity is increased in FXS, without any difference in the temporal dynamics or the shape of the gamma activity. This can be described as a linear increase in gamma power. Further, the relationship between event features and average gamma power does not differ across diagnostic groups, supporting the conclusion that the total amount of gamma activity is increased in FXS, but not the temporal dynamics.

### Absolute event power associated with clinical measures of obsessive compulsive and stereotypic behaviors

Exploratory analysis of correlations between spectral event features and clinical measures indicated that increased gamma event power in the left temporal region was significantly correlated with increased impairment in two clinical measures of obsessive/compulsive behavior and stereotypy, which replicates existing studies showing that measures of behavioral problems are associated with increased gamma single trial power Ethridge et al., 2019). Asynchronous gamma activity has been characterized as cortical noise (Pedapati et al., 2022, Preprint) which could contribute to impaired overall function.

### Correlations between gamma event power and ITC

While gamma event power was associated with decreased gamma synchrony to the chirp stimulus in TDC (see **Table 4**, **Figure 8**), this correlation was not present for subjects with FXS. This aligns with previous existing findings which utilized averaged, non-spectral event, data to show that asynchronous average gamma power was negatively correlated with gamma ITC to the chirp stimulus, with a weaker relationship in FXS (Ethridge et al., 2017, Ethridge et al., 2019). This could be due to a floor effect in subjects with FXS, whose ability to synchronize to the gamma frequency of the chirp stimulus is known to be diminished compared to TDC (Ethridge et al., 2017, Ethridge et al., 2019).

**Table 5 -.** Clinical correlations with absolute event power in left temporal source

| **Event Feature** | **Clinical Measure** | **cor** | **statistic** | **p** | **method** | **p.adj** |
| --- | --- | --- | --- | --- | --- | --- |
| Event power absolute | ADAMS Obsessive/ Compliance Behavior | 0.63 | 1,857 | 0.0002 | Spearman | 0.011 |
| Event power absolute | ABC FXS subscale 3: stereotypy | 0.57 | 1,923 | 0.0010 | Spearman | 0.031 |
| Event power absolute | ABC FXS subscale 5: Inappropriate speech | 0.50 | 2,250 | 0.0050 | Spearman | 0.107 |
| Event power absolute | SCQ Total | 0.46 | 2,699 | 0.0100 | Spearman | 0.162 |
| Event power absolute | ABC FXS subscale 1: irritability/aggression | 0.43 | 2,554 | 0.0172 | Spearman | 0.216 |
| Event power absolute | ABC FXS subscale 4: Hyperactivity/Noncompliance | 0.42 | 2,594 | 0.0199 | Spearman | 0.216 |

### Potential cellular mechanisms of alteration in absolute event power in FXS

The present findings have implications for understanding the circuit level mechanisms of gamma activity in FXS. Increased synchrony between layers 2/3 and layer 5 of the auditory cortex has been hypothesized to contribute to elevated gamma power in FXS (Goswami et al. 2019, Ethridge et al. 2019), which could potentially lead to an increase in event power. The finding of a linear increase in gamma activity in FXS, without changes to the temporal dynamics, constrains the possible circuit and cell level explanations for an increase in gamma power.

**Figure 8 -.**
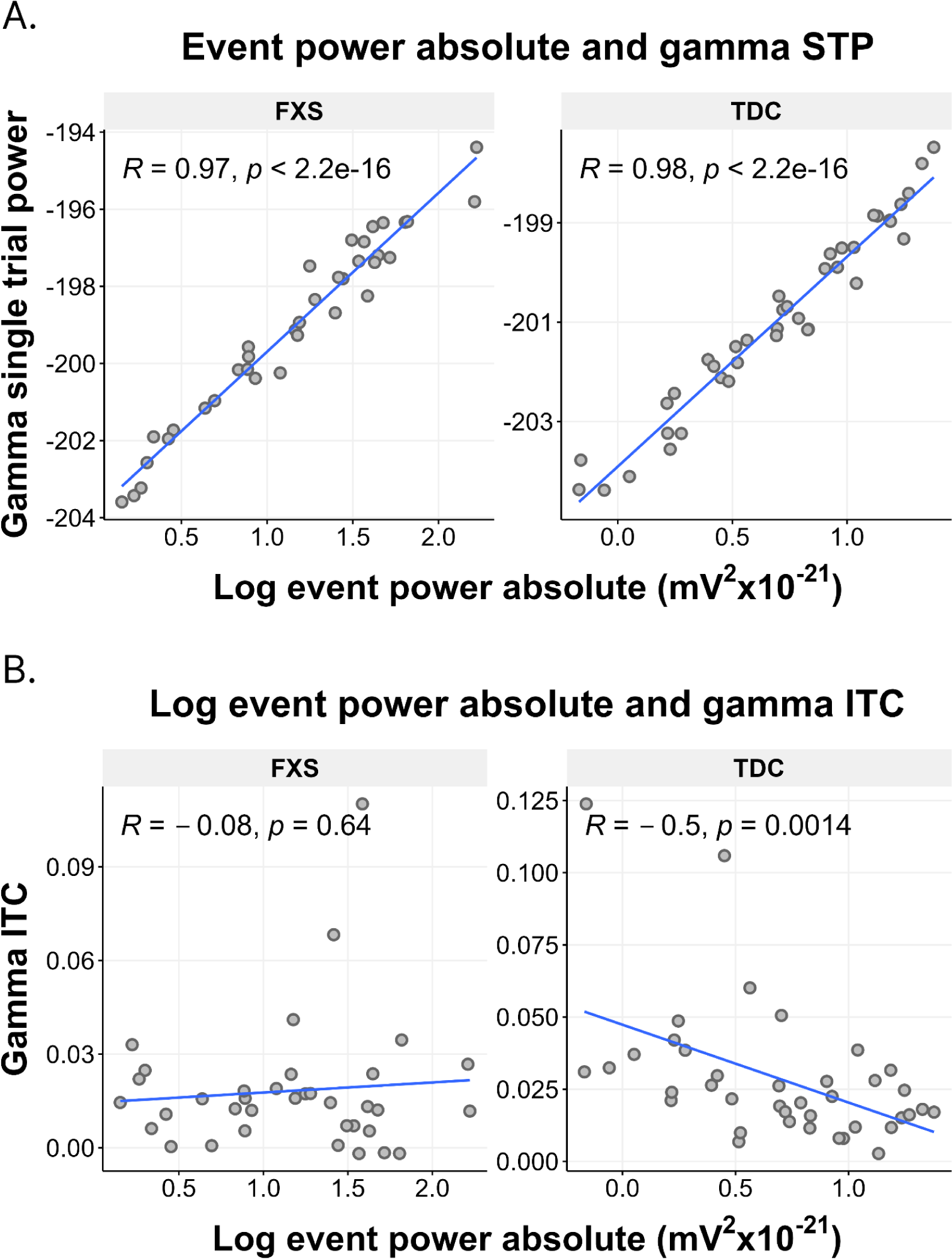
Absolute event power is positively associated with asynchronous gamma power and negatively associated with gamma ITC in TDC. **Figure 8. Absolute event power is positively associated with asynchronous gamma power and negatively associated with gamma ITC in TDC.** Gamma event power in the left temporal source was significantly correlated with gamma STP across all regions (see **Table 3**), Absolute gamma event power was negatively correlated with gamma ITC in typically developing controls (r(39)=-0.5, p=0.0014, adj. p=0.03), but not subjects with FXS (r(36)=-0.08, p=0.64)

### Limitations and future directions

Several limitations must be taken into account when interpreting the present results. First, the cellular mechanisms underlying the observed changes in absolute event power are not fully understood. Further research analyzing gamma spectral events in mouse and circuit models of FXS could allow for a greater understanding of how these changes manifest in neural circuits and lead to. Second, the present findings are limited to an auditory chirp paradigm. Gamma power has been shown to be increased in male subjects with FXS during resting state (Wang et al. 2017, Smith et al., 2021) and other auditory tasks (Ethridge 2017, Ethridge 2019). As the current analysis only considers EEG during auditory chirp, it is unclear whether the current results are specific to auditory stimulation or generalizable to other tasks. Finally, the present study compares averages using nonparametric tests without incorporating other variables into a statistical model, which could be used to take into account variables such as trial count and age to ensure that they do not have interactions with event feature variables of interest. Future studies could analyze gamma spectral events during resting state and other non-auditory EEG paradigms. Finally, future analysis could also be conducted to analyze high power events in the alpha and theta frequency bands, which have been shown to be affected in FXS.

## Conclusions

The present analysis of discrete high power spectral events in subjects with FXS provides insight into the trial level patterns in gamma activity that are hidden in traditional analyses of EEG band power. The lack of differences in the features of spectral events between subjects with FXS and typically developing controls suggests that the circuit level changes in FXS lead to a linear increase in gamma activity without affecting its temporal dynamics. These findings have implications for translational research about gamma activity in FXS.

## Data Availability

All data used in the present study are available upon reasonable request to the authors

