## Supplemental Materials for "Gamma spectral event power is elevated in Fragile X Syndrome and associated with single trial gamma power during auditory chirp"

Supplemental Table 1 - Group comparisons of event feature trial power correlations

| Region | Event feature | n | Statistic | df | p | Method | p.adj |
|---|---|---|---|---|---|---|---|
| RT | Event rate | 75 | 12.477 | 3 | 0.006 | Kruskal-Wallis | 0.236 |
| LF | Event power normalized | 75 | 7.749 | 3 | 0.051 | Kruskal-Wallis | 0.378 |
| LT | Event power normalized | 75 | 7.117 | 3 | 0.068 | Kruskal-Wallis | 0.378 |
| RT | Event power normalized | 75 | 6.859 | 3 | 0.076 | Kruskal-Wallis | 0.378 |
| RT | Event duration | 75 | 6.344 | 3 | 0.096 | Kruskal-Wallis | 0.378 |
| LT | Event power absolute | 75 | 6.331 | 3 | 0.097 | Kruskal-Wallis | 0.378 |
| LO | Event duration | 75 | 6.267 | 3 | 0.099 | Kruskal-Wallis | 0.378 |
| RT | Event power absolute | 75 | 6.241 | 3 | 0.100 | Kruskal-Wallis | 0.378 |
| LP | Event duration | 75 | 6.176 | 3 | 0.103 | Kruskal-Wallis | 0.378 |
| LF | Event duration | 75 | 6.170 | 3 | 0.104 | Kruskal-Wallis | 0.378 |
| RO | Event duration | 75 | 6.170 | 3 | 0.104 | Kruskal-Wallis | 0.378 |
| LT | Event rate | 75 | 5.896 | 3 | 0.117 | Kruskal-Wallis | 0.390 |
| LT | Event fspan | 75 | 5.307 | 3 | 0.151 | Kruskal-Wallis | 0.464 |
| LT | Event duration | 75 | 4.895 | 3 | 0.180 | Kruskal-Wallis | 0.464 |
| RF | Event power absolute | 75 | 4.748 | 3 | 0.191 | Kruskal-Wallis | 0.464 |
| LF | Event rate | 75 | 4.672 | 3 | 0.197 | Kruskal-Wallis | 0.464 |
| LF | Event power absolute | 75 | 4.612 | 3 | 0.202 | Kruskal-Wallis | 0.464 |
| RT | Event fspan | 75 | 4.540 | 3 | 0.209 | Kruskal-Wallis | 0.464 |
| LF | Event fspan | 75 | 4.326 | 3 | 0.228 | Kruskal-Wallis | 0.480 |
| LO | Event power normalized | 75 | 4.143 | 3 | 0.246 | Kruskal-Wallis | 0.492 |
| RF | Event power normalized | 75 | 3.927 | 3 | 0.269 | Kruskal-Wallis | 0.505 |
| LO | Event rate | 75 | 3.852 | 3 | 0.278 | Kruskal-Wallis | 0.505 |
| RP | Event rate | 75 | 3.681 | 3 | 0.298 | Kruskal-Wallis | 0.518 |

| | | | | | | | |
|---|---|---|---|---|---|---|---|
| RF | Event duration | 75 | 3.559 | 3 | 0.313 | Kruskal-Wallis | 0.520 |
| LO | Event power absolute | 75 | 3.418 | 3 | 0.332 | Kruskal-Wallis | 0.520 |
| RF | Event rate | 75 | 3.365 | 3 | 0.339 | Kruskal-Wallis | 0.520 |
| LO | Event fspan | 75 | 3.183 | 3 | 0.364 | Kruskal-Wallis | 0.520 |
| RO | Event rate | 75 | 3.185 | 3 | 0.364 | Kruskal-Wallis | 0.520 |
| LP | Event fspan | 75 | 2.998 | 3 | 0.392 | Kruskal-Wallis | 0.541 |
| LP | Event rate | 75 | 2.575 | 3 | 0.462 | Kruskal-Wallis | 0.616 |
| RO | Event power normalized | 75 | 2.327 | 3 | 0.507 | Kruskal-Wallis | 0.654 |
| RF | Event fspan | 75 | 2.181 | 3 | 0.536 | Kruskal-Wallis | 0.661 |
| LP | Event power normalized | 75 | 2.134 | 3 | 0.545 | Kruskal-Wallis | 0.661 |
| RP | Event fspan | 75 | 1.559 | 3 | 0.669 | Kruskal-Wallis | 0.787 |
| RP | Event power absolute | 75 | 1.417 | 3 | 0.702 | Kruskal-Wallis | 0.787 |
| RO | Event power absolute | 75 | 1.389 | 3 | 0.708 | Kruskal-Wallis | 0.787 |
| LP | Event power absolute | 75 | 1.171 | 3 | 0.760 | Kruskal-Wallis | 0.822 |
| RP | Event power normalized | 75 | 0.821 | 3 | 0.844 | Kruskal-Wallis | 0.888 |
| RO | Event fspan | 75 | 0.694 | 3 | 0.875 | Kruskal-Wallis | 0.897 |
| RP | Event duration | 75 | 0.532 | 3 | 0.912 | Kruskal-Wallis | 0.912 |

Supplemental Figure 1 - Source localized regions

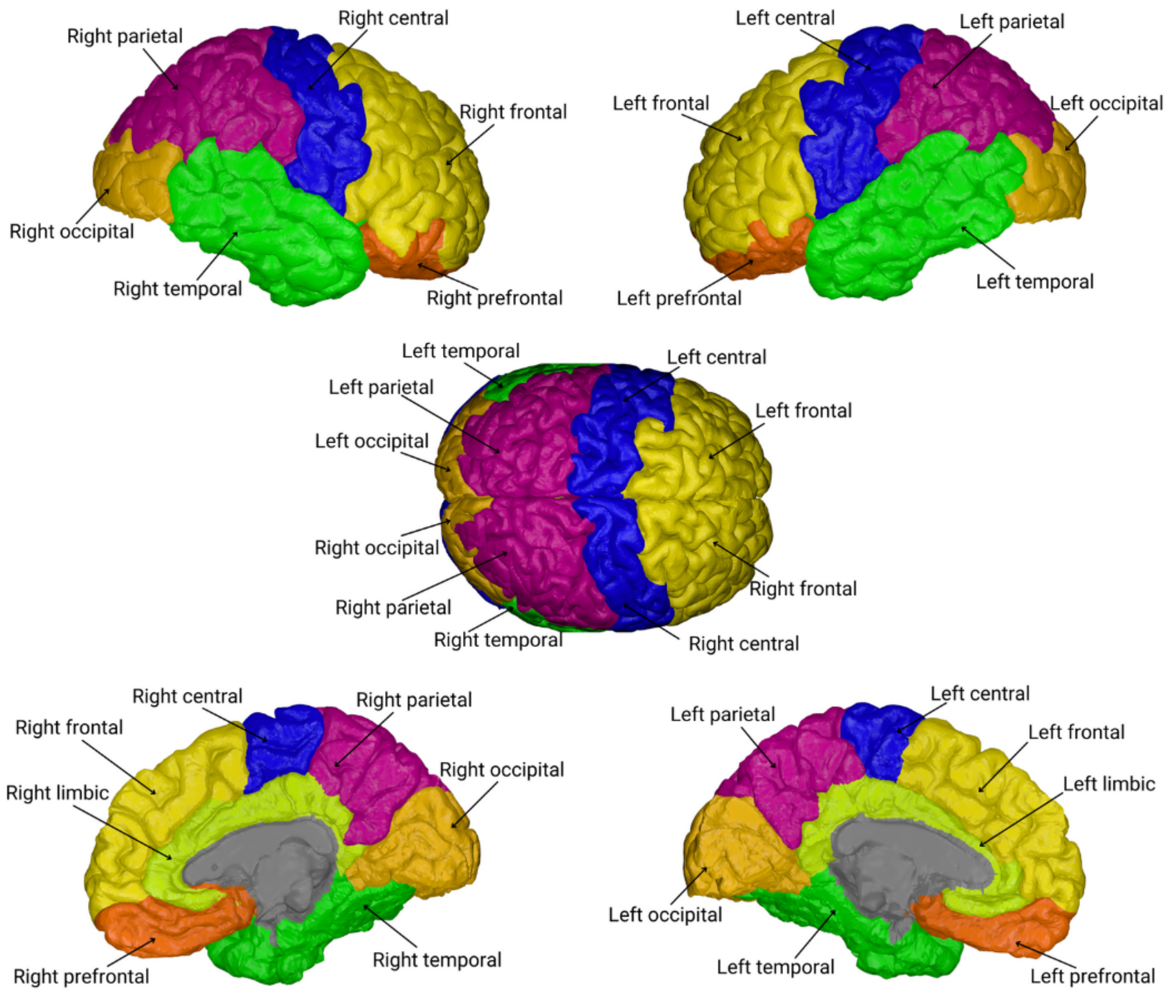

Supplemental Figure 2 - Unchanged normalized event power suggests a linear increase in gamma band activity in FXS.

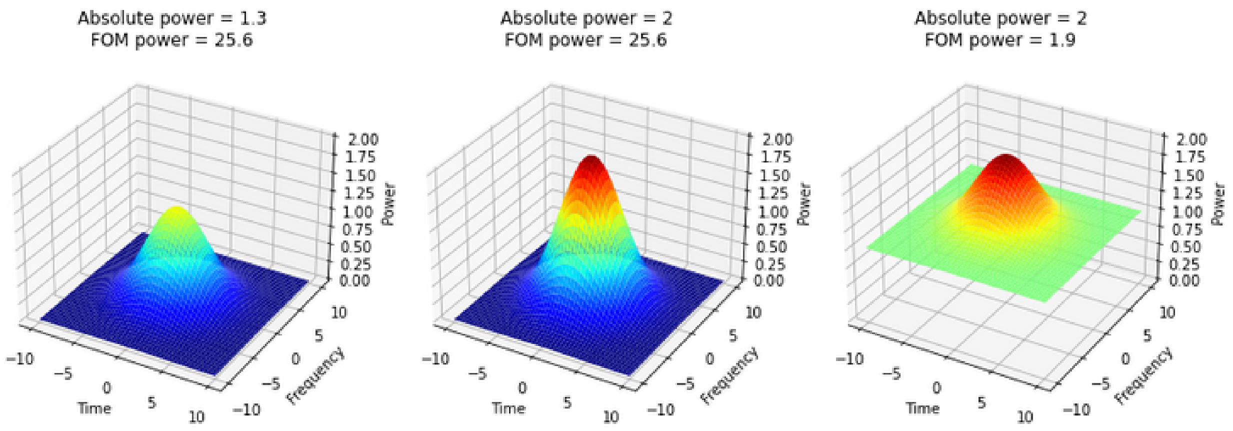


**Supplemental Figure 2. Unchanged normalized event power suggests a linear increase in gamma band activity in FXS.** An increase in event peak power can result from a “linear” increase to the power across the entire trial (panel B) or an “additive” increase in power (panel C) which also affects the normalized (FOM) peak power. The lack of difference in normalized event peak power in FXS suggests that the increase in gamma band activity in FXS does not result from an increase in baseline power outside of high power spectral events, but that the event power is elevated multiplicatively.
